# Pediatric Diabetic Ketoacidosis (PDKA) among newly diagnosed diabetic patients at Dilla University Hospital, Dilla, Ethiopia: prevalence and predictors

**DOI:** 10.1101/2024.03.10.24303986

**Authors:** Dinberu oyamo

## Abstract

**Background:** Diabetic ketoacidosis is a morbid complication of diabetes mellitus, and its occurrence at diagnosis has rarely been studied in Ethiopia, despite the many cases seen in the pediatric population.

**Objective:** To know the prevalence of diabetic ketoacidosis (DKA) among patients with newly diagnosed diabetes mellitus and identify avoidable risk factors.

**Method:** This institution-based retrospective cross-sectional study was conducted from December 25, 2018 to December 25, 2022. Newly diagnosed type1 diabetes mellitus (DM) patients with age < 15 years were included in the study. DKA was diagnosed based on clinical presentation and blood glucose and urine ketone levels. A data collection form was prepared to collect sociodemographic and clinical data. Descriptive, bivariate, and multivariate logistic regression analyses were performed to identify the risk factors.

**Result:** Among the admitted 61 newly diagnosed T1DM pediatric patients, DKA was the first presentation in 37 patients making 60.7% of newly diagnosed T1DM. Mean age at diagnosis was 8(±3.85) years and females were affected more. Clinical presentation revealed vomiting accompanied by signs of dehydration (32.4%), with polysymptoms (29.7%) being the most common. Infectious morbidity occurred in 26 patients, 21 of whom were in the DKA group. Inadequate knowledge of signs and symptoms of DM adjusted odds ratio (AOR=0.07(0.019-0.0897), absence of a family history of DM (AOR=0.129 (0.019-0.897), and presence of infection prior to diagnosis of DKA (AOR=11.69(1.34-10.1) were potential predictors for the development of DKA among newly diagnosed T1DM patients

**Conclusion:** A very high number of children present with DKA at the initial diagnosis of diabetes mellitus (DM), which is attributed to inadequate knowledge of the signs and symptoms of DM and the masking effect of concomitant infections in these children. Healthcare professionals should endeavor to suspect and screen children. Continuous awareness creation of DM at the health professional and community levels is encouraged to diagnose diabetes mellitus earlier and to decrease the prevalence of DKA as an initial presentation.

## Introduction

Diabetic ketoacidosis (DKA) is a common complication of type 1 diabetes mellitus (T1DM) in both children and adolescents ^(1, 2)^.Worldwide, the reported frequencies of DKA as an initial presentation of type 1DM across countries and continents are diverse (as low as 12% and as high as 80%), indicating wide geographic variations in its incidence ^(3)^.

This condition remains a public health concern in sub-Saharan African countries including Ethiopia^(4)^.Small number of reports from Africa have reported that up to 95% of children present with DKA at the time of type 1DM diagnosis^(3,5,6)^; and in Ethiopia, there were two reports showing its high prevalence^(7,8)^.

Earlier diagnosis of diabetes mellitus (DM) in children and adolescents will reduce the number of children presenting with DKA. However, several studies have demonstrated and linked an increased incidence to multiple factors. For instance, younger age ^(9-14)^, uneducated parents, no relative with type1DM, residing in rural areas, and having low economic status were incriminated ^(9, 12, 13, 15)^.Nonetheless, not all the factors were explanatory ^(16)^.

Considering the fewer reports in Ethiopia, the current study was conducted to know the prevalence of DKA among newly diagnosed type 1DM children and to uncover potential factors for its occurrence as a first presentation.

## Methodology

### Study area

The study was conducted at Dilla University hospital, which is located in Dilla town, southern Ethiopia, 360 km away from the Ethiopian capital city, Addis Ababa. The hospital is organized into different departments, and the pediatric and child health departments have emergency,inpatient,outpatient, and neonatal units. The pediatric ward has 61 beds and every month, an average of 875 pediatric patients are admitted for inpatient treatment services.

### Study design, patient selection and data collection technique

The study was conducted using an institution-based retrospective cross-sectional method over a 4-year period, from December 2018 to December 2022. All newly diagnosed type 1 diabetic patients (T1DM) with age less than 15 years old were included in the study and DKA percentage was calculated. An inpatient registration book was used to identify newly diagnosed patients with T1DM, and individual patient charts were retrieved from the card room for data collection. A structured data collection form was prepared to collect data from individual patient charts, and parents were also contacted for data collection. The collected data included socio-demographics (age, sex, family income, parental employment status, parental educational status, and parental marital status), parental knowledge of the signs and symptoms of DM, family history of DM, signs and symptoms (such as polyuria, polydipsia, and weight loss), and the presence of preceding infection.

Diabetic ketoacidosis (DKA) is considered when random blood sugar (RBS) is ≥ 250 mg/dl and ≥ +2 ketones are present in the urine together with dehydration signs and symptom, vomiting,kussmaul respiration and altered mentation. No blood ketone or gas analysis was performed during the study period.

Type 1 DM was diagnosed when the child has classic sympoms of DM plus plasma random blood glucose level of ≥200mg/dl or fasting plasma glucose level of ≥126mg/dl .There was no HbA1C test during the study period.

### Data analysis

Data were checked for completeness and entered into Epi Info v7 and SPSS version 2.1 Data are reported as mean, median, and standard deviation for continuous variables and counts and percentages for categorical variables. The associations between independent variables (age, sex, family income, family history of DM, infection/acute febrile illness before presentation, parental employment, parental educational status, and knowledge of signs and symptoms of DM) and the dependent variable(DKA in newly diagnosed type 1 DM) were assessed using binary logistic regression analysis. Variables that showed an association with the outcome variable (p value of <0.25) were selected for multivariate analysis to control for potential confounders. Adjusted odds ratio (AOR) and 95%confidence interval (CI) were estimated to assess the strength of association. Statistical significance was set at P value of <0.05.

## Result

During the study period (four years), 61 newly diagnosed type 1DM pediatric patients were admitted for inpatient treatment. Diabetic ketoacidosis (DKA) was present in 37 of the 61(60.7%) patients. Of the 61 children, more than half (60.7%) of the patients were females, with a male-to-female ratio of 1:1.54**(Figure 1)**. It was observed that female genders (n= 24, 64.8%) presented with DKA more frequently than did men (n = 13, 35.2%). Both sexes had similar age distributions, with a mean age of 8(±3.85) years. Regarding the marital status of the child parents/guardians of the 61 children, 53(86.9%) were married, five (8.2%) were single, one (1.6%) was widowed, and the rest were divorced. Majority of parents (n=24, 39.3%) earn above 3600 birr (USD),where the majority of the parents/guardians were either employed as civil servants or daily laborers (4(6.6%) unemployed,22(36.1%) civil servants,17(27.9%) daily laborers and other 12 (19.7) have multiple sources of income). Educational levels of the mothers showed that 19(31.1%) were in grade 1-8, 19(31.1%) can read and write, 16(26.2%) were in grade 8-12 and only 5 (8.2%) of them were above grade 12. Similarly paternal education level showed 20(32.8%) were above grade12, 16(26.2%) were in grade 9-12, 14(23%) were in grade 1-8 and 11(18.2%) can only read and write. **(Table 1)**.

**Table 1.**
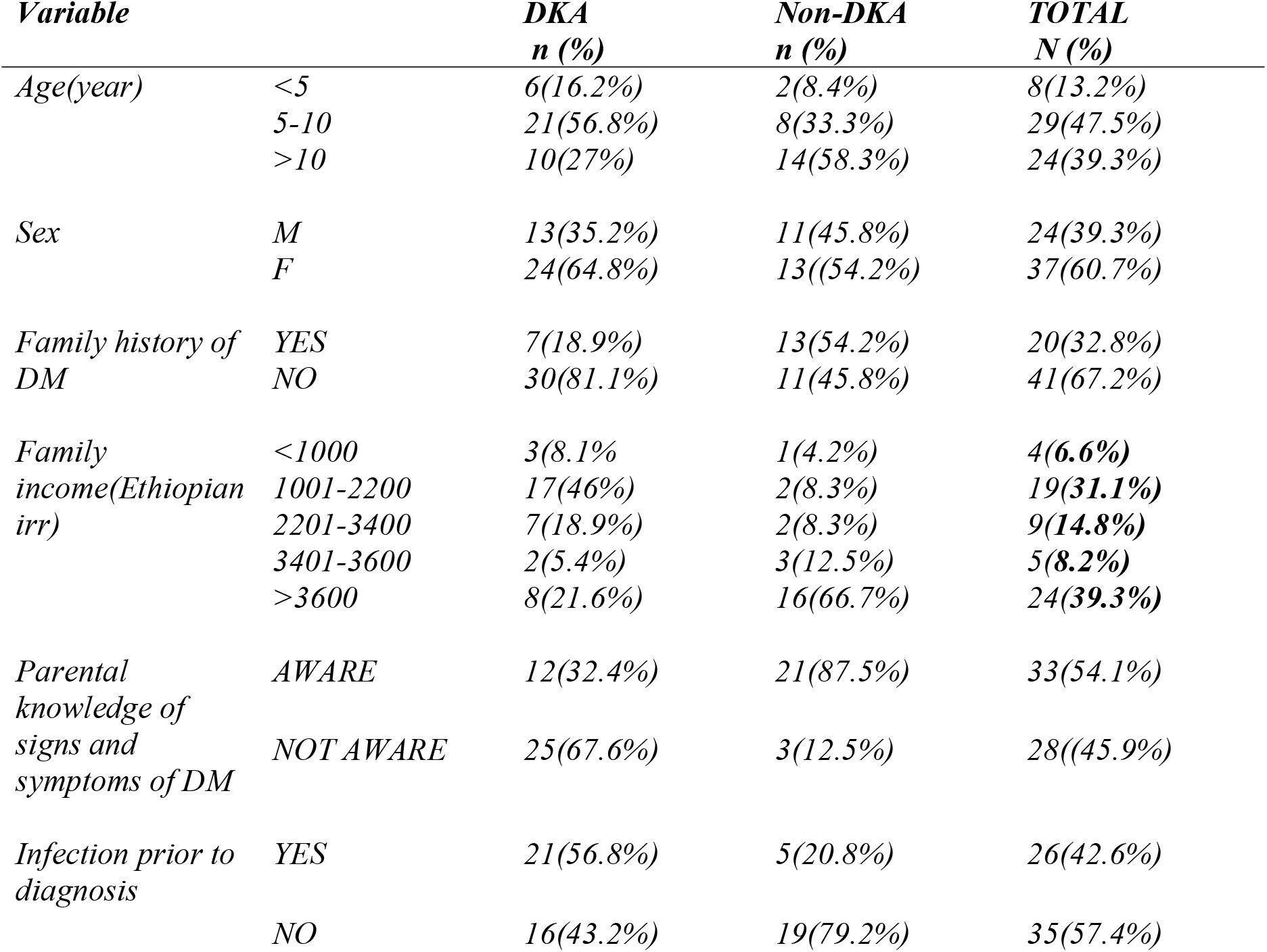
socio-demographic characteristics of newly diagnosed T1DM with DKA and without DKA.

| **Variable** |  | **DKA**<br>**n (%)** | **Non-DKA**<br>**n (%)** | **TOTAL**<br>**N (%)** |
| --- | --- | --- | --- | --- |
| Age(year) | <5 | 6(16.2%) | 2(8.4%) | 8(13.2%) |
|  | 5-10 | 21(56.8%) | 8(33.3%) | 29(47.5%) |
|  | >10 | 10(27%) | 14(58.3%) | 24(39.3%) |
| Sex | M | 13(35.2%) | 11(45.8%) | 24(39.3%) |
|  | F | 24(64.8%) | 13((54.2%) | 37(60.7%) |
| Family history of DM | YES | 7(18.9%) | 13(54.2%) | 20(32.8%) |
|  | NO | 30(81.1%) | 11(45.8%) | 41(67.2%) |
| Family income(Ethiopian irr) | <1000 | 3(8.1% | 1(4.2%) | 4(**6.6%**) |
|  | 1001-2200 | 17(46%) | 2(8.3%) | 19(**31.1%**) |
|  | 2201-3400 | 7(18.9%) | 2(8.3%) | 9(**14.8%**) |
|  | 3401-3600 | 2(5.4%) | 3(12.5%) | 5(**8.2%**) |
|  | >3600 | 8(21.6%) | 16(66.7%) | 24(**39.3%**) |
| Parental knowledge of signs and symptoms of DM | AWARE | 12(32.4%) | 21(87.5%) | 33(54.1%) |
|  | NOT AWARE | 25(67.6%) | 3(12.5%) | 28((45.9%) |
| Infection prior to diagnosis | YES | 21(56.8%) | 5(20.8%) | 26(42.6%) |
|  | NO | 16(43.2%) | 19(79.2%) | 35(57.4%) |

**Figure 1.**
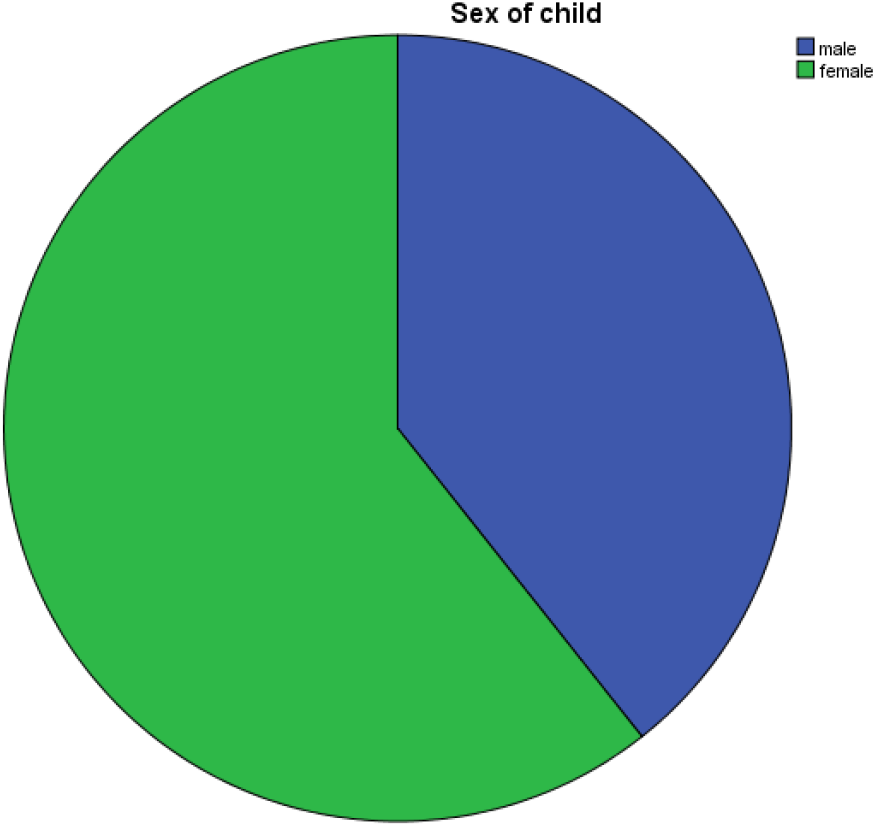
Sex distribution of newly diagnosed T1DM.

The majority, 41 (67.2%),of the children did not have a family history of diabetes, and 33(54%) parents knew the signs and symptoms of DM/DKA. The clinical presentations of the 37 children with DKA were abdominal pain in 10(27.2%), loss of consciousness in 4(10.8%), vomiting accompanied by dehydration in 12(32.4%), and polysymptoms in the remaining 11(29.7%). Infection occurred in 26 children, 21 of whom had DKA. Of the 21 infections in DKA, pneumonia was the most commonly diagnosed infection in seven (33.3%) children and diarrheal disease in seven (33.3%), followed by urinary tract infection in four (19%), and acute tonsillar pharyngitis in three (14.3%). **(Table 1, Table 2)**

On bivariate analysis, family history of DM, p = 0.006,OR=0.193 CI(0.063-0.623),family income value 0.00,OR=2.26(1.437-3.55),parents’ knowledge of signs and symptoms, p value 0.00,OR =0.069 CI(0.017-0.276), and preceding infection, OR=4.987 CI(1.532-16.23) showed significant associations and were candidates for multivariate analysis.Female sex (p = 0.64) and age group of 5.1-10 years (p = 0.733, OR=0.89 CI (0.455-1.739) frequently presented with DKA, however the association was not significant. Similarly, parental educational level, marital status, occupation and signs, and symptoms of DM/DKA that the child had prior to diagnosis did not have a significant association.

**Table 2:** clinical presentations of newly diagnosed DM with DKA.

| *Clinical presentations* | *N (%)* |
| --- | --- |
| *Abdominal pain* | *10(27.2%)* |
| *Loss of consciousness* | *4(10.8%)* |
| *Vomiting* | *12(32.4%)* |
| *Poly symptom* | *11(29.7%)* |
| *Infectious morbidity(N=21)* |  |
| *Pneumonia* | *7(33.3%)* |
| *Diarrhea* | *7(33.3%)* |
| *UTI* | *4(19%)* |
| *Tonsilopharyngitis* | *3(14.3%)* |

In multivariate analysis, variables that showed a significant association were also strongly associated in multivariate analysis too. Those children whose parents/guardians knew the signs and symptoms of DKA/DM were 93% less likely to develop DKA at the time of the initial diagnosis of DM (p = 0.017; AOR=0.070(0.008-0.618). Children who had a first-degree relative with DM had a significant association with the diagnosis of DKA at the initial presentation of DM, with a p-value of 0.039 AOR=0.129(0.019-0.897). New-onset DM with DKA are 2 times higher in children with a family monthly income of >3600birr (64.87 USD) than in those with a monthly income of < 1000 birr (18.02 USD). **(Table 3)**

**Table 3.**
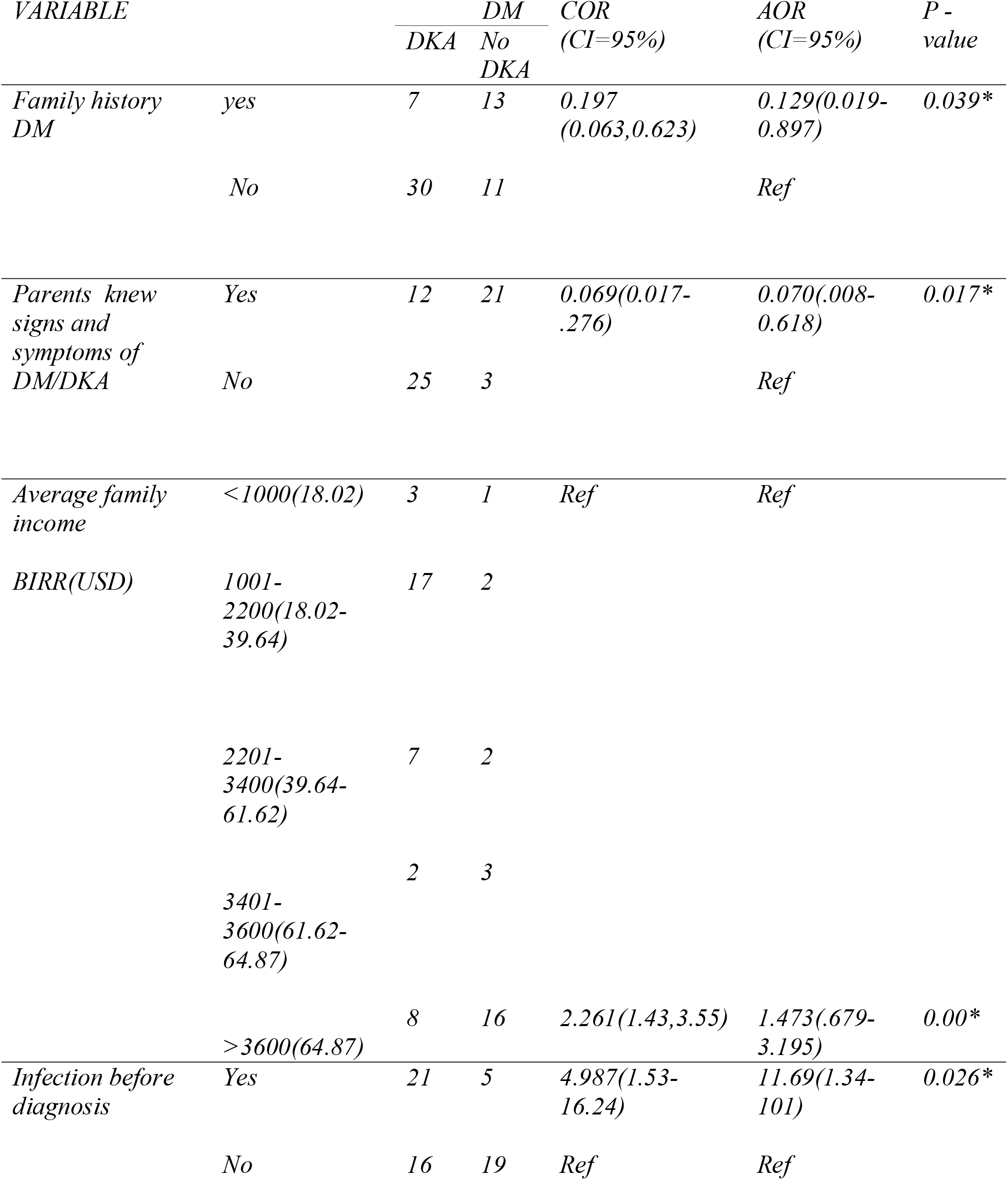
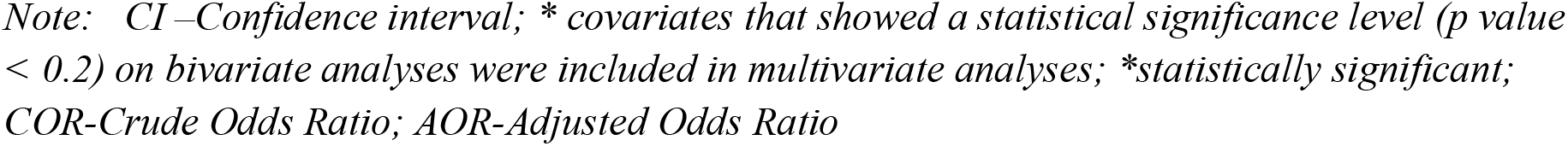
Bivariable and multivariable analysis of newly diagnosed T1DM.

| VARIABLE |  | DM |  | COR | AOR | P - |
| --- | --- | --- | --- | --- | --- | --- |
|  |  | DKA | No DKA | (CI=95%) | (CI=95%) | value |
| Family history DM | yes | 7 | 13 | 0.197<br>(0.063,0.623) | 0.129(0.019-0.897) | 0.039* |
|  | No | 30 | 11 |  | Ref |  |
| Parents knew signs and symptoms of DM/DKA | Yes | 12 | 21 | 0.069(0.017-.276) | 0.070(.008-0.618) | 0.017* |
|  | No | 25 | 3 |  | Ref |  |
| Average family income | <1000(18.02) | 3 | 1 | Ref | Ref |  |
| BIRR(USD) | 1001-2200(18.02-39.64) | 17 | 2 |  |  |  |
|  | 2201-3400(39.64-61.62) | 7 | 2 |  |  |  |
|  | 3401-3600(61.62-64.87) | 2 | 3 |  |  |  |
|  | >3600(64.87) | 8 | 16 | 2.261(1.43,3.55) | 1.473(.679-3.195) | 0.00* |
| Infection before diagnosis | Yes | 21 | 5 | 4.987(1.53-16.24) | 11.69(1.34-101) | 0.026* |
|  | No | 16 | 19 | Ref | Ref |  |

*Note: CI—Confidence interval; \* covariates that showed a statistical significance level (p value < 0.2) on bivariate analyses were included in multivariate analyses; \*statistically significant; COR-Crude Odds Ratio; AOR-Adjusted Odds Ratio*

## Discussion

The frequency of childhood and adolescent diabetic ketoacidosis (DKA) as an initial presentation of diabetes mellitus varies worldwide. This retrospective study aimed to review the number of children presenting with DKA at diagnosis and to reveal the background risks for its occurrence in rural and resource-limited settings in Ethiopia. In this study, 60.7% of the children had DKA as an initial presentation of diabetes mellitus. This is much higher than the study conducted in Ethiopian capital, Addis Ababa (35.8%), which was 80% 15 years back^(7,17^), but much lower than in northern part of Ethiopia, Tigray(78.7%)^(8)^.The high level literacy of the community and the availability of wide range of awareness creation methods(such as screening,posters,flyers and awareness campaigns) in the capital might explain the sharp decline in the incidence. The model was demonstrated in Germany, where the incidence was reduced from 28% to 16% through the Stuttgart ketoacidosis campaign, which focused on the symptoms of type 1 DM ^(10)^.In addition, the current prevalence is higher than that in other African countries such as Egypt(46%)^(15)^,Sudan(17.6%)^(18)^, and Tanzania(43%)^(19)^,but lower than that in two studies in Nigeria(100%)^(9)^ (77%)^(20)^.Outside the African continent, the reported prevalence was UK(25%)^(21)^,Germany(20%)^(22)^,Finland(19. 4%)^(23)^,Sweden(16%), USA (30%), Canada (18.6%)^(16)^,Kuwait (35.9%), and China (50.1%)^(24)^, all showing wide geographic variation in its incidence. Various factors have been identified for a wide range of variations, including variations in the prevalence of diabetes, demonstrating an inverse relationship between type 1 diabetes mellitus prevalence and DKA incidence ^(25, 26)^.

The diagnosis of type 1 DM at a mean age of 8 years and the frequent occurrence of DKA at a younger age in the current study agree with several previous studies conducted in different setups^(11,12,13,19,25)^.In contrast to earlier studies in Ethiopia^(7,8,17)^,Nigeria^(7),^Saudi Arabia ^(12)^ and wales^(27)^, the current study observed a higher number of female children presenting with DKA as an initial presentation of type 1 DM than males. This observation is in line with other studies ^(28)^; however, none of the previous studies could explain the increased occurrence in female gender. A recent study from England, Germany, Wales, Austria, and the United States failed to explain female predominance in terms of clinical, biological, and physiological factors ^(29)^.

The finding of vomiting accompanied by dehydration signs, polysymptoms, and abdominal pain in the present study aligns with studies done in northern and central Ethiopia^(7,8,17,30)^ as well as other studies where polysymptoms were the dominant presentations^(3,19,24,31,32,33,34)^.In addition, the identification of infectious precipitants, particularly pneumonia, diarrheal diseases, and urinary tract infections, and as a predictor of DKA also favor the hypothesis that infections lead to acute metabolic decompensation and insulin resistance and subsequent development of diabetic ketoacidosis^(28)^.

One prominent finding of the current study was that children whose parents who were unaware of the signs and symptoms of DM/DKA were more likely to develop DKA as an initial presentation. Moreover, the study also found that having no parental family history of diabetes and a high monthly family income led to a diagnosis of DKA at presentation. This finding is in line with studies that have suggested that creating awareness of the signs and symptoms of classic symptoms of DM reduces the incidence of DKA at diagnosis^(3, 31, 35-40)^.Different strategies have been used to create awareness in the community, such as awareness campaigns on classic symptoms of DM/DKA at the local (such as schools, parents/care givers, and health care workers) or nationwide levels. Such methods are effective across countries ^(10, 38, 41, 42, 43)^ however, some are not ^(44, 45)^.

In agreement with earlier studies^(7)^ and Germany^(46)^, and despite most parents/guardians having primary and secondary education levels, the current study demonstrate that education levels of parents doesn’t have impact on the prevalence of DKA at presentation; however, several studies have demonstrated a protective effect^(28, 47-50)^.The lack of awareness as well as the lack of information provided on pediatric diabetes or DKA contributed to the insignificant effect of educational level on occurrence of DKA. Similarly, in concordance with other studies ^(51, 52, 53)^.The current study found no association between parental employment and marital status and DKA as an initial presentation.

### Limitations

Despite the potential limitations (retrospective nature and lost charts and torn pages), the study showed areas that need to be addressed promptly.

## Conclusions

In conclusion, the current study indicates that the number of children presenting with DKA at the time of type 1 DM diagnosis was very high compared to both national and global studies. The unpredictable course in younger children, masking effect of infectious comorbidities, and low awareness of type 1 DM at the individual level were the main factors. Effective awareness creation methods must be implemented at both the health professional and community levels to reduce the burden.

## Data Availability

available upon reasonable request to the authors

## Abreviations

DM: DM-diabetes mellitus
T1DM: type 1 DM
DKA: diabetic ketoacidosis
USD: united states dollar.

## Ethical clearance and consent to participate

Before conducing the research, ethical approval was obtained from Dilla university institutional review board on December 2022. Verbal consent was obtained to collect data from parents/guardians, after explaining the purpose of the research and ensuring strict confidentiality. Permission was obtained from Dilla university hospital to collect data from patient charts. Data were collected based on declaration of Helsinki.

## Funding

None

## Competing interests

None

## Acknowledgement

I would like to thank all parents and guardians who were willing to provide their consent and participate in the research. I also acknowledge healthcare workers at the Department of Pediatrics and Child Health, who provided unreserved care for these children.

## Notes

### Competing Interest Statement

The authors have declared no competing interest.

### Funding Statement

This study did not receive any funding

### Summary of Updates

This revision only made to make formatting the manuscript.no other changes were made.

## References

1. Wolfsdorf J, Craig ME, Daneman D, et al. Diabetic ketoacidosis. Pediatr. Diabetes. .2007;8:28–43

2. Jefferies C, Cutfield SW, Derraik JG, et al. 15-year incidence of diabetic ketoacidosis at onset of type 1 diabetes in children from a regional setting (Auckland, New Zealand). Sci. Rep. 2015; 5:1–7. doi:10.1038/srep10358).

3. Usher Smith JA, Thompson MJ, Ercole A, Walter FM. Variation between countries in the frequency of diabetic ketoacidosis at first presentation of type 1 diabetes in children: a systematic review. Diabetologia. 2012; 55:2878–2894. doi:10.1007/s00125-012-2690-2

4. Non-Communicable O. National strategic action plan (nsap) for prevention & control (of non-communicable diseases in Ethiopia. (2016), Google Scholar. Ministry of Health

5. Murunga AN, Owira PM. Diabetic ketoacidosis an overlooked child killer in sub-Saharan Africa. Tropical Med Int Health. 2013;18:1357–1364

6. Smith CP. Diabetic Ketoacidosis, Current Pediatrics.2006; 16,111–116.

7. Fantahun B, Gedlu E. Prevalence of diabetic ketoacidosis in newly diagnosed diabetes mellitus pediatric patients in TikurAnbessa Specialized Hospital. Off Organ Ethiopian Pediatr Soc. 2008;4:1

8. Hadgu FB, Sibhat GG, Gebretsadik LG. Diabetic ketoacidosis in children and adolescents with newly diagnosed type 1 diabetes in Tigray, Ethiopia: retrospective observational study. Pediatric Health Med Ther. 2019; 10:49–55. Published 2019 May 23. doi:10.2147/PHMT.S207165

9. Umar UI, Muhammed IL, Aliyu I. Retrospective review of presentation of newly diagnosed children with diabetes mellitus in a Nigerian rural setting. Med J DY Patil Vidyapeeth.2019; 12:490–4.

10. Holder M, Ehehalt S. Significant reduction of ketoacidosis at diabetes onset in children and adolescents with type 1 diabetes–The Stuttgart Diabetes Awareness Campaign, Germany. Pediatr Diabetes.2020; 21:1227–31. doi:10.1111/pedi.13064

11. Al Rashed A. Pattern of presentation in type 1 diabetic patients at the diabetes center of a university hospital. Ann Saudi Med. 2011; 31: 243.

12. Abdullah MA. Epidemiology of type I diabetes mellitus among Arab children. Saudi Med J. 2019;26: 911–917

13. Chowdhury S .Puberty and type 1 diabetes. Indian J Endocrinol Metab.2015;19(Suppl 1): S51-S54.doi:10.4103/2230-8210.155402

14. Todorović S. Milenković T, Mitrović K, Plavšić L, Panić Zarić S, Vuković R. High prevalence of diabetic ketoacidosis in children with newly diagnosed type 1 diabetes in Belgrade, Serbia: 10-year tertiary centre experience. Zdravstvena zaštita. 2019; 48(4):7–14. doi:10.5937/ZZ1904007T

15. Noha Arafa,mona Mamdouh Hassan,Shimaa atef,Asmaa salah eldin fathallah,amany Ibrahim. Clinical characterstics and precipitating factor(s) associated with diabetic ketoacidosis presentation in children with newly diagnosed diabetes. Clinical diabetology.2020; 9(5):286–292. doi:10.5603/DK.2020.0042

16. J. A. Usher-Smith, M. Thompson, A. Ercole, and F. M. Walter. Variation between countries in the frequency of diabetic ketoacidosis at first presentation of type 1 diabetes in children: a systematic review.Diabetologia.2012; 55(11): 2878–2894.

17. Atkilt HS, Turago MG, Tegegne BS. Clinical characteristics of diabetic ketoacidosis in children with newly diagnosed type 1 diabetes in Ethiopia. A cross-sectional study. PLoS One. 2017; 12(1). doi:10.1371/journal.pone.0169666

18. Ahmed AM, Khabour OF, Ahmed SM, Alebaid IA, Ibrahim AM. Frequency and severity of ketoacidosis at diagnosis among childhood type 1 diabetes in Khartoum state, Sudan. Afri Health Sci. 2020; 20(2): 841–848.10.4314/ahs.v20i2.38)

19. Kipasika H, Majaliwa E, Kamala B, Mungai L. Clinical Presentation and Factors Associated with Diabetic Ketoacidosis at the Onset of Type-1 Diabetes Mellitus in Children and Adolescent at Muhimbili National Hospital,Tanzania:A cross sectional study. int J Diabetes Clin Res.2020;7:126. 10.23937/2377-3634/1410126

20. N. Onyiriuka and E. Ifebi.Ketoacidosis at diagnosis of type 1 diabetes in children and adolescents: frequency and clinical characteristics. Journal of Diabetes and Metabolic Disorders. 2013; 12(1):47.

21. K. Lokulo-Sodipe, R. J. Moon, J. A. Edge, and J. H. Davies.Identifying targets to reduce the incidence of diabetic ketoacidosis at diagnosis of type 1 diabetes in the UK.Archives of Disease in Childhood. 2014; 99(5):438–442.

22. Segerer H, Wurm W, Grimsmann JM, et al. Diabetic ketoacidosis at manifestation of type 1 diabetes in childhood and adolescence— incidence and risk factors. Dtsch Arztebl Int .2021; 118: 367–72. doi:10.3238/arztebl.m2021.0133

23. Hekkala A, Reunanen A, Koski M, Knip M, Veijola R; Finnish Pediatric Diabetes Register. Age related differences in the frequency of ketoacidosis at diagnosis of type 1 diabetes in children and adolescents. Diabetes Care. 2010; 33:1500–1502.

24. Peng W, Yuan J, Chiavaroli V, et al. 10-Year Incidence of Diabetic Ketoacidosis at Type 1 Diabetes Diagnosis in Children Aged Less Than 16 Years From a Large Regional Center (Hangzhou, China). Front. Endocrinol. 2021; 12:653519. doi:10.3389/fendo.2021.653519

25. Cherubini V, Grimsmann JM, Åkesson K, et al. Temporal trends in diabetic ketoacidosis at diagnosis of paediatric type 1 diabetes between 2006 and 2016: results from 13 countries in three continents. Diabetologia. 63(8):1530–1541. 2020; doi:10.1007/s00125-020-05152-1

26. Große J, Hornstein H, Manuwald U, Kugler J, Glauche I, Rothe U. Incidence of diabetic ketoacidosis of new-onset type 1 diabetes in children and adolescents in different countries correlates with Human Development Index (HDI): an updated systematic review, meta-analysis, and meta-regression. Horm Metab Res. 2018; 50: 209–2210.1007/s00125-020-05152-1

27. Patterson CC, Harjutsalo V, Rosenbauer J et al. Trends and cyclical variation in the incidence of childhood type 1 diabetes in 26 European centres in the 25 year period 1989-2013: a multicenter prospective registration study. Diabetologia.2019; 62(3):408–417. 10.1007/s00125-018-4763-3

28. Usher-Smith JA, Thompson MJ, Sharp SJ, and Walter FM. Factors associated with the presence of diabetic ketoacidosis at diagnosis of diabetes in children and young adults: a systematic review. BMJ. 2011; 343:d4092. doi:10.1136/bmj.d4092

29. Maahs DM, Hermann JM, Holman N et al. Rates of diabetic ketoacidosis: international comparison with 49,859 pediatric patients with type 1 diabetes from England, Wales, the US, Austria, and Germany. Diabetes Care.2015; 38:1876–1882

30. Teshome D. Diabetic Ketoacidosis in an Addis Abeba Children’s Hospital. Ethiop Med J. 1992; 30:7–11

31. Lévy-Marchal C, Patterson CC, Green A; EURODIAB ACE Study Group. Europe and Diabetes. Geographical variation of presentation at diagnosis of type I diabetes in children: the EURODIAB study. European and Dibetes. Diabetologia. 2001; 44 Suppl 3:B75–B80. doi:10.1007/pl00295800

32. Souza LCVF, Kraemer GC, Koliski A, et al. DIABETIC KETOACIDOSIS AS THE INITIAL PRESENTATION OF TYPE 1 DIABETES IN CHILDREN AND ADOLESCENTS: EPIDEMIOLOGICAL STUDY IN SOUTHERN BRAZIL. Rev Paul Pediatr. 2019; 38:e2018204. Published 2019 doi:10.1590/1984-0462/2020/38/2018204Nov25.

33. Roche EF, Menon A, Gill D, Hoey H. Clinical presentation of type 1 diabetes. Pediatr Diabetes. 2005; 6(2):75–78. doi:10.1111/j.1399-543X.2005.00110.x

34. Noha Arafa,mona Mamdouh Hassan,Shimaa atef,Asmaa salah eldin fathallah,amany Ibrahim. Clinical characterstics and precipitating factor(s) associated with diabetic ketoacidosis presentation in children with newly diagnosed diabetes. Clinical diabetology.2020; 9(5):286–292. doi:10.5603/DK.2020.0042

35. Bui H, To T, Stein R, Fung K, Daneman D. Is diabetic ketoacidosis at disease onset a result of missed diagnosis? J Pediatr.2010; 156:472–7.

36. Barker JM, Goehrig SH, Barriga K, et al, DAISY study. Clinical characteristics of children diagnosed with type 1 diabetes through intensive screening and follow-up. Diabetes Care. 2004; 27(6): 1399–1404.10.2337/diacare.27.6.1399

37. Vanelli M, Chiari G, Ghizzoni L, Costi G, Giacalone T, Chiarelli F. Effectiveness of a prevention program for diabetic ketoacidosis in children. Diabetes Care. 1999; 22:7–9. 10.2337/diacare.22.1.7

38. Vanelli M, Chiari G, Lacava S, Iovane B. Campaign for diabetic ketoacidosis prevention still effective 8 years later. Diabetes Care. 2007; 30:e12. 10.2337/dc07-0059

39. Triolo TM, Chase HP, Barker JM, the DPT-1 Study Group .Diabetic subjects diagnosed through the diabetes prevention trial type 1 (DPT-1) are often asymptomatic with normal A1C at diabetes onset. Diabetes Care .2009; 32(5):769–773.10.2337/dc08-1872

40. Elding Larsson H, Vehik K, Bell R, et al, TEDDY Study Group., SEARCH Study Group., Swediabkids Study Group., DPV Study Group., Finnish Diabetes Registry Study Group. Reduced prevalence of diabetic ketoacidosis at diagnosis of type 1 diabetes in young children participating in longitudinal follow-up. Diabetes Care .2011; 34(11):2347–2352.10.2337/dc11-1026

41. Rabbone I, Maltoni G, Tinti D, et al. Diabetic ketoacidosis at the onset of disease during a national awareness campaign: a 2-year observational study in children aged 0-18 years. Arch Dis Child .2020; 105:363–6. doi:10.1136/archdischild2019-316903

42. King BR, Howard NJ, Verge CF, et al. A diabetes awareness campaign prevents diabetic ketoacidosis in children at their initial presentation with type 1 diabetes. Pediatr Diabetes. 2012; 13:647–51. 10.1111/j.1399-5448.2012.00896.x

43. Lansdown AJ, Barton J, Warner J, et al. Prevalence of ketoacidosis at diagnosis of childhood onset Type 1 diabetes in Wales from 1991 to 2009 and effect of publicity campaign. Diabetes Med .2012; 29:1506–9. doi:10.1111/j.1464-5491.2012.03638.x

44. Derraik JGB, Cutfield WS, Maessen SE et al. A brief campaign to prevent diabetic ketoacidosis in children newly diagnosed with type 1 diabetes mellitus: the NO-DKA study. Pediatr Diabetes. 2018; 19(7):1257–1262.10.1111/pedi.12722

45. Fritsch M, Schober E, Rami-Merhar B, Hofer S, Fröhlich-Reiterer E, Waldhoer T. Diabetic ketoacidosis at diagnosis in Austrian children: a population-based analysis, 1989-2011. J Pediatr .2013; 163:1484.e1– 8.e1. doi:10.1016/j.jpeds.2013.06.033

46. Rosenbauer J, Icks A, Giani G. Clinical characteristics and predictors of severe ketoacidosis at onset of type 1 diabetes mellitus in children in a North Rhine-Westphalian region, Germany. J Pediatr Endocrinol Metab.2002;15:1137–45

47. Sadauskaite-Kuehne V, Samuelsson U, Jasinskiene E, et al. Severity at onset of childhood type 1 diabetes in countries with high and low incidence of the condition. Diabetes Res Clin Pract .2002; 55:247–54.

48. Komulainen J, Lounamaa R, Knip M, Kaprio EA, Akerblom HK. Ketoacidosis at the diagnosis of type 1 (insulin dependent) diabetes mellitus is related to poor residual beta cell function. Arch Dis Childhood.1996; 75:410–5.

49. Rewers A, Klingensmith G, Davis C, et al. Presence of diabetic ketoacidosis at diagnosis of diabetes mellitus in youth: The search for diabetes in youth study. Pediatrics.2008; 121: e1258–e1266.

50. Komulainen J, Kulmala P, Savola K, et al. Clinical, autoimmune, and genetic characteristics of very young children with type 1 diabetes. Childhood Diabetes in Finland (DiMe) Study Group. Diabetes Care.1999; 22: 1950–1955.

51. Smith CP, Firth D, Bennett S, Howard C, Chisholm P. Ketoacidosis occurring in newly diagnosed and established diabetic children. Acta Paediatrica.1998;87:537–41

52. Hamilton DV, Mundia SS, Lister J. Mode of presentation of juvenile diabetes. BMJ .1976; 2:211–2.

53. Sadauskaite-Kuehne V, Samuelsson U, Jasinskiene E, et al. Severity at onset of childhood type 1 diabetes in countries with high and low incidence of the condition. Diabetes Res Clin Pract. 2002;55:247–54

